# Progress and Inequality in Diabetes Care Cascade in Indonesia: A National Health Survey Analysis (2013–2023)

**DOI:** 10.64898/2026.07.29.26359228

**Authors:** Farizal R. Muharram, Risyad A. Siregar, M.Q.B. Zulfikar, Aqsha Nur, Indah Suci Widyahening, Goodarz Danaei

## Abstract

**Background:** To examine trends in Indonesia’s diabetes care cascade from 2013 to 2023, identify key determinants, and assess progress toward global targets of 80% diagnosis and 80% glycemic control among those diagnosed.

**Methods:** We analyzed nationally representative data from Indonesia’s Health Surveys in 2013, 2018, and 2023. Diabetes was defined using fasting plasma glucose and oral glucose tolerance tests. We estimated diagnosis, treatment, and control rates and examined sociodemographic predictors of cascade progression using survey-weighted logistic regression models.

**Results:** Between 2013 and 2023, the prevalence of diabetes among adults aged ≥15 years remained stable, ranging from 10.7% to 11.8%. Diagnosis increased from 15.1% (95% CI: 13.4–16.7) to 20.7% (18.5–22.9), treatment nearly doubled from 10.5% (9.1–11.9) to 19.0% (16.9–21.2), and control rose modestly from 4.6% (3.6–5.6) to 6.5% (5.2–7.8). Older age, urban residence, higher socioeconomic status, and insurance coverage were associated with greater progression through the cascade. Wealth-related inequalities persisted in 2023: one-third of cases were diagnosed among the richest (35.3% [29.0–41.7]) versus only 11.0% (7.9–14.2%) among the poorest. Compared with the lowest quintile, wealthier individuals had higher odds of diagnosis (AOR 3.55 [2.11–5.98] for diagnosis, 3.59 [2.01– 6.41] for treatment, and 2.24 [1.12–4.51] for control).

**Conclusions:** Indonesia achieved meaningful improvements in the diabetes care cascade over the past decade, yet remains far below global 80/80 targets, with nearly 80% of cases undiagnosed and control below 10%. Persistent wealth-based inequities highlight that near-universal insurance coverage has not been translated into equitable care access, underscoring the need for equity-focused screening and primary care strengthening.

**Twitter Summary:** Indonesia has improved diabetes diagnosis and treatment since 2013, but glycemic control remains low and unequal, especially by wealth and rurality. Targeted screening + quality care are urgently needed.

**ARTICLE HIGHLIGHTS:**

- Why did we undertake this study? We aimed to evaluate Indonesia’s progress in diabetes management from 2013 to 2023 and to identify sociodemographic inequalities affecting the care cascade, in line with global 80-80 targets.
- What is the specific question(s) we wanted to answer? We asked how diagnosis, treatment, and control rates changed over time and which factors including age, residence, socioeconomic status, and insurance—predict better cascade progression.
- What did we find? Diabetes prevalence remained stable at 10–12%. Diagnosis rose from 15% to 21%, treatment nearly doubled, but control increased only slightly. About 80% of adults remained undiagnosed, with pronounced wealth-related disparities.
- What are the implications of our findings? Findings highlight diagnosis as the main bottleneck and call for equity-focused strategies, stronger primary care, and expanded screening and insurance coverage.

## INTRODUCTION

Indonesia is the world’s fourth most populous country, with approximately 276 million people distributed across more than 17,000 islands.^1^ The national diabetes prevalence in Indonesia is projected to rise from 9.2% in 2020 to 16.1% by 2045^2^. adding more than 23 million people with diabetes within two decades. To reduce this burden, WHO Member States, including Indonesia, adopted five global diabetes coverage targets at the 2022 World Health Assembly^3^. These include 80% diagnosis and 80% glycemic control among diagnosed. Prior studies estimated that around 70% of people with diabetes in Indonesia remained undiagnosed^4,5^, with only 30.8% of treated individuals reaching glycemic control.^6^ Indonesia’s diabetes policies are set out in the National Medium-Term Development Plan and the Ministry of Health Strategic Plan (2020–2024^7^, with screening devolved to district governments under the Minimum Service Standards (SPM)^8^. Screening is delivered through primary care and community Posbindu posts using capillary blood glucose tests. Diagnosed cases are covered by national health insurance (JKN).

Combined with a decentralized health system, this geography creates substantial heterogeneity in healthcare access. Launched in January 2014, Indonesia’s national health insurance (JKN) expanded rapidly to become the world’s largest single-payer scheme, reaching approximately 267 million members by 2023 (about 95% of the population). However, near-universal enrolment has not translated into equitable use: roughly half of members do not access outpatient benefits, with lower use concentrated among informal-sector workers in lower wealth quintiles. The JKN also established a Chronic Disease Management Program (Prolanis) in primary care in 2014.^9^ Medications are also provided free of charge at primary care facilities, though availability may vary.^10^ Despite these policies, diabetes care performance remains poor. Key barriers include limited diagnostic capacity in rural and remote facilities, low health literacy and awareness, particularly among lower-income populations, and persistent geographic and financial barriers to accessing benefits. ^4,5,7,8^

Using three rounds of Indonesia’s national health surveys (2013, 2018, 2023), we examine trends in diabetes diagnosis, treatment, and control, and identify the sociodemographic determinants of cascade progression. Findings aim to inform equity-focused screening and primary-care strengthening aligned with global diabetes targets.

## METHODS

### Data Source and Population

This study utilized nationally representative health surveys conducted by the Indonesian government: Indonesian Basic Health Research (*Riskesdas*) 2013 and 2018, and the Indonesian Health Survey (SKI) 2023. These cross-sectional surveys employed a stratified, multistage, systematic random sampling design based on census blocks to ensure national representativeness. Details of the sampling methodology are provided in **Supplementary Text 1 and Supplementary Table 1**. The study population included all respondents aged ≥15 years randomly selected from the subsample of the National Health Survey. We included data on past diabetes status (diagnosis and treatment), laboratory tests, including fasting plasma glucose (FPG) and oral glucose tolerance test (OGTT) results. In addition, HbA1c data were analyzed for 2023, as they were unavailable in the previous survey. Pregnant individuals were excluded. Missing covariate data were categorized as "missing". Characteristics of excluded and missing participants are provided in Supplementary Table 7. Details on the glucose test protocol were provided in **Supplementary Text 2**. To ensure national representativeness, we constructed calibrated person-level weights for each survey round, aligning the biomedical subsample with the full household survey across age (5-year groups, capped at ≥80), sex, urban/rural residence, and the proportion diagnosed with diabetes. Calibration used iterative proportional fitting (raking). Reweighting balanced the survey and laboratory samples; detailed results are in Supplementary Table 6.

### Outcomes and Covariates

The primary outcomes were diabetes prevalence and the diabetes care cascade (diagnosis, treatment, and control). Prevalence was determined using a combined approach capturing diagnosed and undiagnosed cases (see Supplementary Tables 2 and 3): individuals were classified as having diabetes if they met at least one of (1) self-reported physician diagnosis, (2) current use of oral diabetes medication or insulin, or (3) positive laboratory results (FPG ≥126 mg/dL or OGTT ≥200 mg/dL). The care cascade comprised three sequential stages, all with diabetes prevalence as the denominator: diagnosed (informed of diabetes by a physician or ever used diabetes medication), treated (diagnosed individuals using medication regularly or occasionally), and controlled (FPG <126 mg/dL or OGTT <180 mg/dL).

Covariates included age (15–39.9, 40–60, >60), sex (male/female), and BMI classified per WHO guidelines as underweight (<18.5 kg/m²), normal (18.5–24.9), overweight (25.0–29.9), and obese (≥30.0). Household wealth was assessed using a 21-item asset-based questionnaire; principal component analysis generated a wealth index for each survey year, used to stratify households into quintiles from poorest (Q1) to richest (Q5). Subdistricts were classified as urban or rural based on government-defined structures.^11^ Details on categorization are shown in **Supplementary Table 4**.

### Statistical Analysis

Descriptive statistics summarized baseline characteristics at each stage of the diabetes care cascade, incorporating survey design weights, clustering, and stratification to ensure national representativeness and correct variance estimation. Progression through the cascade stages (diagnosis, treatment, glycemic control) was examined using design-based, population-averaged logistic regression, modeling each stage separately with survey year as the predictor. Models adjusted for sex, age, BMI, wealth quintile, and insurance status, with adjusted odds ratios (AORs) and 95% CIs reported. To assess whether improvements varied by socioeconomic group, interaction terms between survey year and wealth quintile were added:

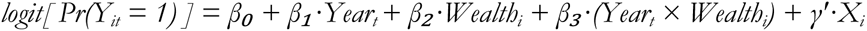

where Y_it is the binary cascade-stage indicator for individual i in round t (2013 reference), Wealth_i is the wealth quintile (Q1 reference), and X_i is a covariate vector (sex, age category, BMI category, insurance status, urban/rural residence). The joint significance of the interaction was tested using a design-based Wald test. Following significant interactions, post hoc marginal analyses were conducted using estimated marginal means and pairwise contrasts to compare wealth quintiles within each year. We pre-specified one primary contrast per outcome × year: the Q1-Q5 difference on the probability scale. A Bonferroni correction across the three rounds within each outcome (α = 0.0167) controlled the family-wise error rate. All analyses used the survey package in R version 4.2. A sensitivity analysis assessed the robustness of the glycemic control definition by applying alternative thresholds (FPG <140 mg/dL, OGTT <200 mg/dL, and HbA1c <6.5%).

### Data and Resource Availability

The microdata is available from Indonesia’s Ministry of Health Data Management Laboratory (layanandata.kemkes.go.id) upon reasonable request and prior written permission. Aggregated outputs from this study are available from the corresponding author.

### Ethical Approval

Ethics and permissions for conducting this study were in accordance with the Ethical Approval for RISKESDAS 2018 from the Ethical Committee of Health Research, NIHRD, Ministry of Health, Republic of Indonesia, No. LB.02.01/2/KE.267/ 2017. As RISKESDAS 2018 allowed the authors to analyze the dataset through the data management laboratory in NIHRD, the ethics referred to the ethical clearance of RISKESDAS 2018

## RESULTS

This study included 68,634 participants from biomedical samples collected in 2013, 2018, and 2023 (**Table 1, Supplementary Table 5**). Diabetes prevalence increased from 10.7% (95% Confidence Interval: 10.2–11.2%) in 2013 to 11.8% (11.3–12.3%) in 2018, then slightly decreased to 11.3% (10.7–11.9%) in 2023. In 2013, 15.1% (13.6–16.7%) of the diabetes population were diagnosed, increasing to 18.6% (17.0–20.3%) in 2018 and 20.7% (18.5–22.9%) in 2023 (**Figure 1**), with the latter number translating to approximately 5.6 million of the 27.6 million people with diabetes being diagnosed.

**Figure 1.**
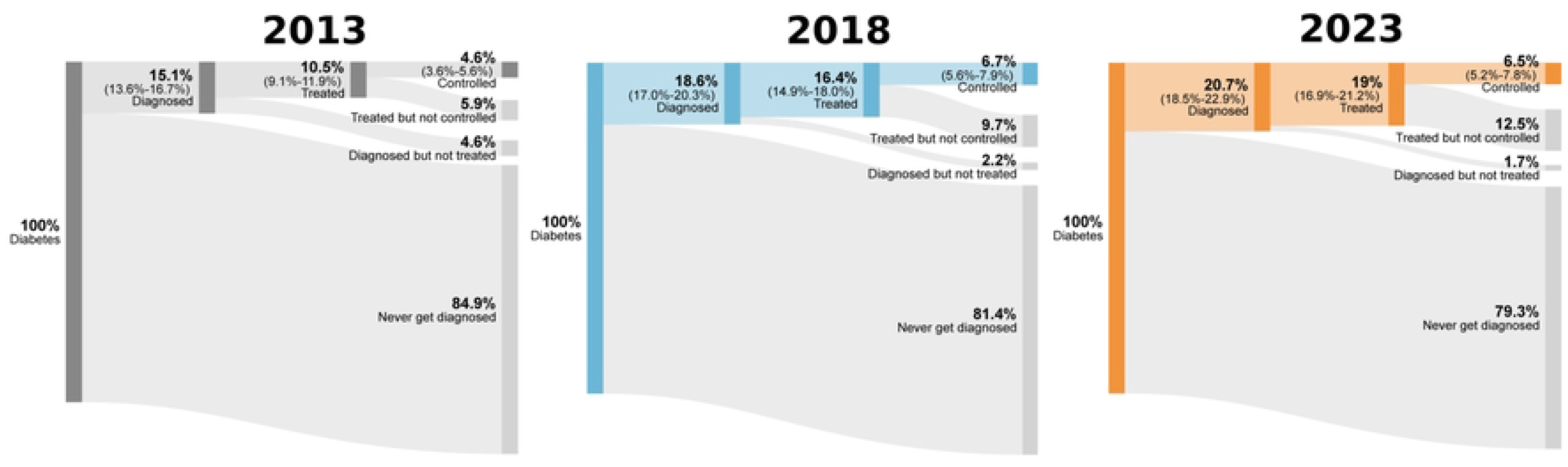
Diabetes Cascade of Care Across Time (2013-2023) Survey-weighted percentages of adults with diabetes achieving each cascade step—diagnosed, treated (any glucose-lowering medication), and controlled—are shown for 2013, 2018, and 2023, with 95% CIs. “Control” follows the study’s prespecified thresholds (see Methods). Estimates reflect nationally representative samples.

**Table 1.** Weighted Sample Characteristics of Indonesia Health Survey 2013-2023.

| Characteristic | 2013 | 2018 | 2023 |
| --- | --- | --- | --- |
| **Diabetes** | 10.7% (10.2-11.2%) | 11.8% (11.3-12.3%) | 11.3% (10.7-11.9%) |
| **Sex** |  |  |  |
| Female | 50.2% (49.3-51.0%) | 50.2% (49.3-51.1%) | 49.7% (48.5-50.9%) |
| Male | 49.8% (49.0-50.7%) | 49.8% (48.9-50.7%) | 50.3% (49.1-51.5%) |
| **Domicile** |  |  |  |
| Rural | 49.0% (48.1-49.9%) | 44.7% (43.8-45.5%) | 41.4% (40.2-42.6%) |
| Urban | 51.0% (50.1-51.9%) | 55.3% (54.5-56.2%) | 58.6% (57.4-59.8%) |
| **Age group** |  |  |  |
| 15-40 | 60.7% (59.9-61.5%) | 56.1% (55.3-57.0%) | 53.8% (52.6-55.0%) |
| 40-60 | 29.6% (28.9-30.4%) | 32.1% (31.3-32.8%) | 32.7% (31.7-33.7%) |
| >60 | 9.7% (9.24-10.1%) | 11.8% (11.3-12.3%) | 13.5% (12.8-14.2%) |
| **BMI Category** |  |  |  |
| <18.5 | 12.9% (12.3-13.5%) | 11.0% (10.4-11.6%) | 9.3% (8.59-10.1%) |
| >=30.0 | 5.4% (5.08-5.77%) | 9.2% (8.74-9.65%) | 9.7% (9.04-10.3%) |
| 18.5-24.9 | 60.8% (59.9-61.6%) | 54.2% (53.3-55.1%) | 52.9% (51.7-54.2%) |
| 25.0--29.9 | 19.6% (19.0-20.3%) | 24.4% (23.7-25.1%) | 25.8% (24.8-26.8%) |
| Missing | 1.3% (1.04-1.68%) | 1.2% (0.90-1.65%) | 2.3% (1.75-2.89%) |
| **Insurance Status** |  |  |  |
| No Insurance | 50.9% (50.0-51.7%) | 35.7% (34.8-36.5%) | 28.1% (26.9-29.2%) |
| Have Insurance | 49.1% (48.3-50.0%) | 64.3% (63.5-65.2%) | 71.9% (70.8-73.1%) |
| **Wealth Quintile** |  |  |  |
| 1 (Poorest) | 14.3% (13.7-14.9%) | 17.2% (16.6-17.9%) | 18.8% (17.9-19.7%) |
| 2 | 18.7% (18.1-19.3%) | 18.5% (17.8-19.2%) | 19.6% (18.7-20.5%) |
| 3 | 22.0% (21.3-22.7%) | 19.3% (18.6-20.1%) | 20.3% (19.4-21.2%) |
| 4 | 24.3% (23.5-25.1%) | 20.6% (19.9-21.3%) | 20.5% (19.5-21.5%) |
| 5 (Wealthiest) | 20.8% (20.0-21.6%) | 24.4% (23.5-25.2%) | 20.9% (19.8-22.0%) |

The proportion of individuals with diabetes receiving treatment also rose over time (**Figure 1**). In 2013, 10.5% (9.1–11.9%) of individuals with diabetes were undergoing treatment, which grew to 16.4% (14.9–18.0%) in 2018 and further to 19.0% (16.9–21.2%) in 2023. This rise in treatment rates occurred alongside increases in diagnosis rates and in the proportion of the diagnosed population that received treatment. For example, the proportion of the population diagnosed and treated was 68.4% (63.6–73.1%) in 2013; by 2023, it had increased to 92.1% (90.1–94.0%) (P < 0.0001). (**Table 2**).

**Table 2.** Proportion of People with Diabetes Who Are Treated Among Those Diagnosed, and Controlled Among Those Treated, 2013–2023.

| Variable | Proportion of Treatment in Diagnosed Diabetes |  |  | Proportion of Controlled in Treated Diabetes |  |  |
| --- | --- | --- | --- | --- | --- | --- |
|  | 2013 | 2018 | 2023 | 2013 | 2018 | 2023 |
| **Overall** | **68.4% (63.6-73.1)** | **87.9% (85.7-90.0)** | **92.1% (90.1-94.0)** | **44.2% (37.2-51.1)** | **40.4% (35.0-45.7)** | **33.9% (28.2-39.6)** |
| **Sex** |  |  |  |  |  |  |
| Female | 67.4% (61.4-73.4) | 89.7% (87.4-91.9) | 91.9% (89.5-94.4) | 36.6% (28.4-44.7) | 36.6% (30.8-42.4) | 32.5% (26.3-38.8) |
| Male | 69.5% (61.9-77.1) | 85.8% (81.9-89.7) | 92.2% (89.2-95.2) | 52.7% (41.3-64.0) | 45.0% (35.8-54.2) | 35.6% (25.4-45.8) |
| **Age Group** |  |  |  |  |  |  |
| 15-40 | 48.1% (36.4-59.9) | 72.4% (61.7-83.0) | 81.1% (71.4-90.9) | 68.0% (52.8-83.2) | 64.4% (45.8-83.1) | 36.7% (15.3-58.1) |
| 40-60 | 76.0% (70.8-81.2) | 90.5% (88.2-92.8) | 93.4% (91.2-95.6) | 36.2% (27.9-44.4) | 34.0% (28.2-39.8) | 31.3% (24.3-38.2) |
| >60 | 73.7% (63.9-83.5) | 91.7% (88.5-94.9) | 95.2% (93.1-97.4) | 46.4% (30.9-61.9) | 42.2% (33.3-51.1) | 36.9% (27.3-46.5) |
| **BMI** |  |  |  |  |  |  |
| <18.5 | 56.9% (37.4-76.4) | 84.8% (75.2-94.5) | 90.8% (83.4-98.3) | 40.4% (15.5-65.3) | 45.2% (27.5-62.9) | 45.7% (19.9-71.6) |
| 18.5-24.9 | 65.5% (58.5-72.4) | 86.9% (83.3-90.4) | 92.1% (89.1-95.1) | 43.8% (34.1-53.5) | 42.1% (33.5-50.7) | 27.9% (20.1-35.7) |
| 25.0--29.9 | 73.4% (65.5-81.4) | 89.1% (85.8-92.4) | 91.3% (88.0-94.6) | 39.9% (27.5-52.2) | 36.5% (28.5-44.4) | 39.1% (28.7-49.4) |
| >=30.0 | 70.7% (57.0-84.5) | 88.6% (84.0-93.2) | 94.1% (90.6-97.7) | 51.5% (30.1-73.0) | 35.4% (22.0-48.8) | 36.0% (23.5-48.6) |
| **Insurance Status** |  |  |  |  |  |  |
| Have Insurance | 70.4% (64.4-76.3) | 88.9% (86.5-91.3) | 93.6% (91.8-95.4) | 40.4% (31.9-48.9) | 44.3% (38.1-50.6) | 34.3% (28.1-40.5) |
| No Insurance | 66.6% (59.2-73.9) | 84.4% (79.7-89.2) | 83.5% (76.1-90.9) | 47.8% (36.9-58.7) | 26.6% (18.1-35.2) | 31.5% (16.9-46.2) |
| **Wealth Quintile** |  |  |  |  |  |  |
| 1 (Poorest) | 33.7% (17.3-50.1) | 77.4% (68.6-86.2) | 89.7% (84.8-94.6) | 50.1% (23.5-76.7) | 43.1% (28.6-57.6) | 44.6% (28.7-60.6) |
| 2 | 52.5% (37.8-67.3) | 81.7% (74.0-89.3) | 92.9% (88.5-97.3) | 41.8% (21.6-61.9) | 43.9% (29.4-58.4) | 38.8% (24.2-53.5) |
| 3 | 67.6% (57.9-77.4) | 87.7% (82.3-93.1) | 91.2% (86.6-95.8) | 47.6% (33.3-61.9) | 42.3% (25.8-58.8) | 28.3% (18.8-37.8) |
| 4 | 78.3% (71.6-85.1) | 89.7% (85.8-93.6) | 90.6% (85.7-95.4) | 48.4% (37.3-59.4) | 29.3% (20.1-38.4) | 32.0% (20.8-43.2) |
| 5 (Wealthiest) | 70.6% (61.6-79.5) | 91.1% (88.2-94.1) | 93.5% (90.4-96.6) | 38.3% (25.2-51.5) | 43.2% (35.3-51.1) | 32.4% (21.4-43.3) |
| **Domicile** |  |  |  |  |  |  |
| Rural | 61.4% (53.6-69.2) | 82.6% (78.1-87.1) | 90.7% (86.9-94.4) | 50.3% (40.2-60.4) | 44.0% (34.9-53.2) | 36.9% (26.7-47.2) |
| Urban | 72.2% (66.4-78.1) | 90.0% (87.6-92.3) | 92.7% (90.5-94.9) | 41.3% (32.3-50.3) | 39.0% (32.6-45.5) | 32.6% (25.7-39.5) |

The proportion of the diabetes population achieving glycemic control increased from 4.6% (3.6-5.6%) in 2013 to 6.5% (5.3-7.7%) in 2018 and remained stagnant at 6.5% (5.2-7.8%) in 2023, representing only 3.39 million of 27.5 million individuals with diabetes achieving control. Using different thresholds to define control did not change the results notably (See **Supplementary Table 8**). Among treated participants, the proportion achieving control was significantly reduced from 44.2% (37.2-51.1%) in 2013 to 33.9% (28.2-39.6%) in 2023 (P=0.026) (**Supplementary Table 14**).

There were no significant differences between males and females across all survey years in the diabetes care cascade (**Supplementary Figure 1**). (**Supplementary Tables 9, 10, 11, and 12**). Urban areas consistently had higher diagnosis rates in all survey years (**Figure 2**). In 2013, the diagnosis rate was 10.8% (9.2-12.5%) in rural areas compared to 19.4% (16.8-22.0%) in urban areas. By 2023, these rates increased to 15.5% (12.6-18.5%) in rural areas and 23.8% (20.9-26.8%) in urban areas (**Figure 2**). Across all years, urban residents had a higher likelihood of diabetes diagnosis (AOR 1.53 (1.32–1.78)), treatment (AOR 1.59 (1.35–1.88)), and control (AOR 1.35 (1.05–1.73)) (**Figure 4**).

**Figure 2.**
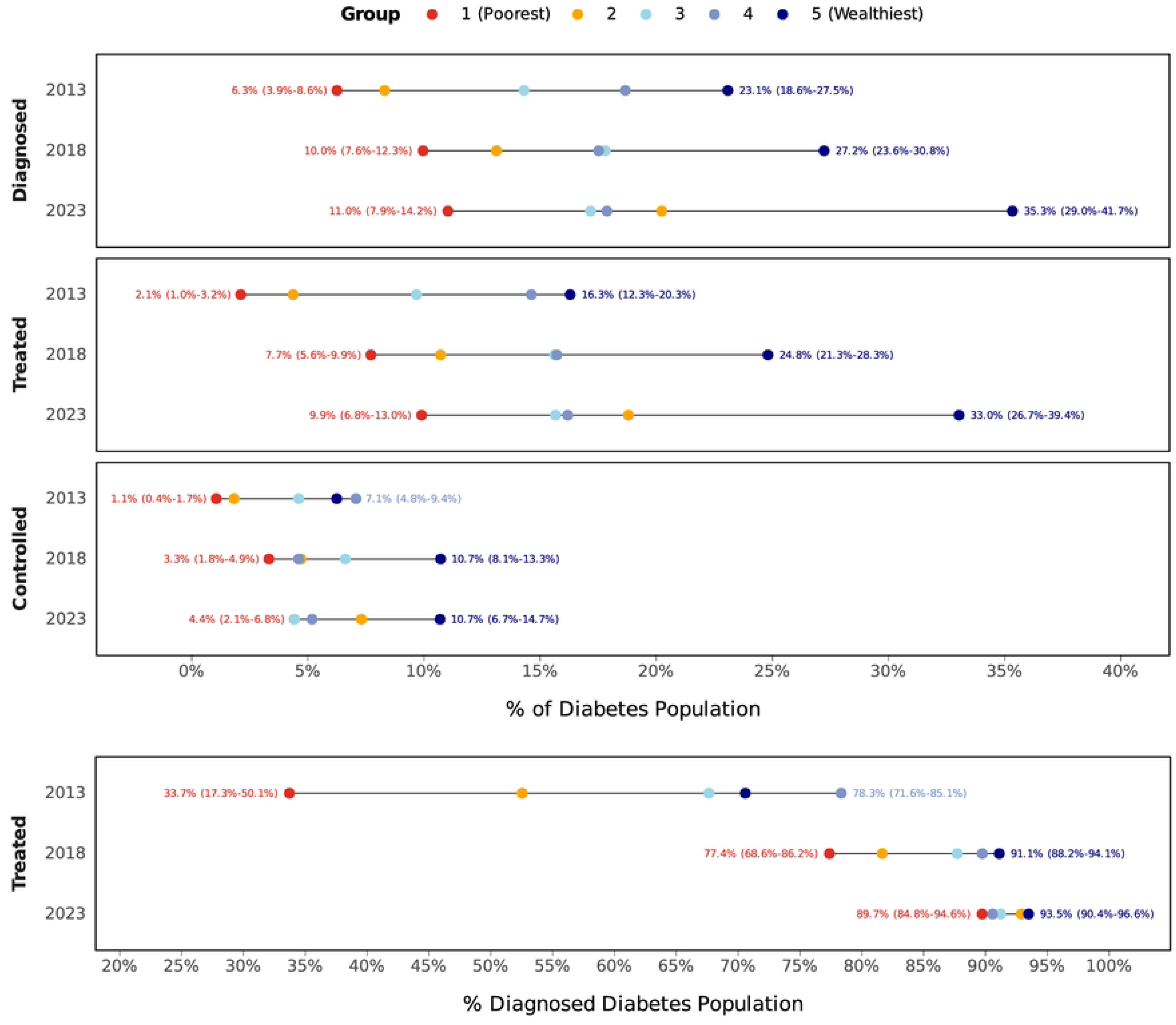
Diabetes Gap of Care Coverage Between Urban and Rural Over Time, National Health Survey 2013-2023.

The improvement in diabetes care varied by age group, with the most significant progress observed among those aged 60 or above. Over the 10 years, this age group showed up to twice the improvement across all stages of the care cascade compared to 2013 as a baseline. The diagnosis rate increased from 13.9% (10.7-17.1%) to 23.1% (19.0-27.1%), the treatment rate rose from 10.2% (7.2-13.2%) to 22.0% (18.0-25.9%), and glycemic control increased from 4.8% (2.7-6.8%) to 8.1% (5.6-10.6%). The 40–60 age group demonstrated moderate improvement, following a similar but less pronounced trend. Meanwhile, the 15–40 age group shows the lowest performance and the least improvement across all stages of the care cascade (**Supplementary Figure 2, Supplementary Tables 10, 11, and 12**). Across BMI categories, normal-weight (18.5–24.9 kg/m²) and overweight (25.0–29.9 kg/m²) individuals had the highest rates of diagnosis, treatment, and glycemic control throughout the care cascade. In contrast, both underweight (<18.5 kg/m²) and obese individuals (≥30.0 kg/m²) showed lower performance at every cascade stage, with underweight individuals having the lowest diagnosis and treatment rates overall (see **Supplementary Tables 10, 11, and 12**). On the other hand, underweight and obese individuals showed a lower care cascade prevalence. Subgroup-by-year rates of glycemic control among treated patients are reported in **Supplementary Table 13**.

Disparities in wealth quintile were shown across the care cascade. In 2023, individuals in the highest wealth quintile (Q5) had the highest rates of diagnosis (Q1 11.0% [7.9-14.2%] vs Q5 35.3% [29.0-41.7%]), treatment (Q1 9.9% [6.8-13.0] vs Q5 33.0% [26.7-39.4]), and glycemic control (Q1 4.4% [2.1-6.8%] vs Q5 10.7% [6.7-14.7]) (**Figure 3**). Adding interaction terms between survey year and wealth quintile significantly improved model fit for all three outcomes (design-based Wald F = 3.89, p = 0.0001 for diagnosis; F = 5.21, p < 0.0001 for treatment; F = 2.66, p = 0.007 for control; Supplementary Table 15). In 2013, individuals in Q5 had significantly higher adjusted odds of being diagnosed (AOR = 5.03, 95% CI 2.93–8.63), treated (AOR = 7.10, 3.80–13.25), and achieving glycaemic control (AOR = 5.34, 2.38–12.00) compared with those in Q1, and all three Q5 vs Q1 contrasts remained significant after Bonferroni adjustment in every round (all p_adj_ ≤ 0.026). By 2023, these gaps had narrowed: the Q5 vs Q1 disparity decreased to AOR 3.55 (2.11–5.98) for diagnosis, 3.59 (2.01–6.41) for treatment, and 2.24 (1.12–4.51) for control — corresponding to roughly 46%, 57%, and 45% reductions in the odds-scale gap, respectively. All other within-year quintile contrasts (the remaining nine pairs per year per outcome) were treated as exploratory and Tukey-HSD-adjusted; complete adjusted p-values are reported in Supplementary Table 16.

**Figure 3.** Care Cascade of Diabetes Between Poorest(Q1) and Richest(Q5) Quintiles Between 2013 and 2023.

**Figure 4.**
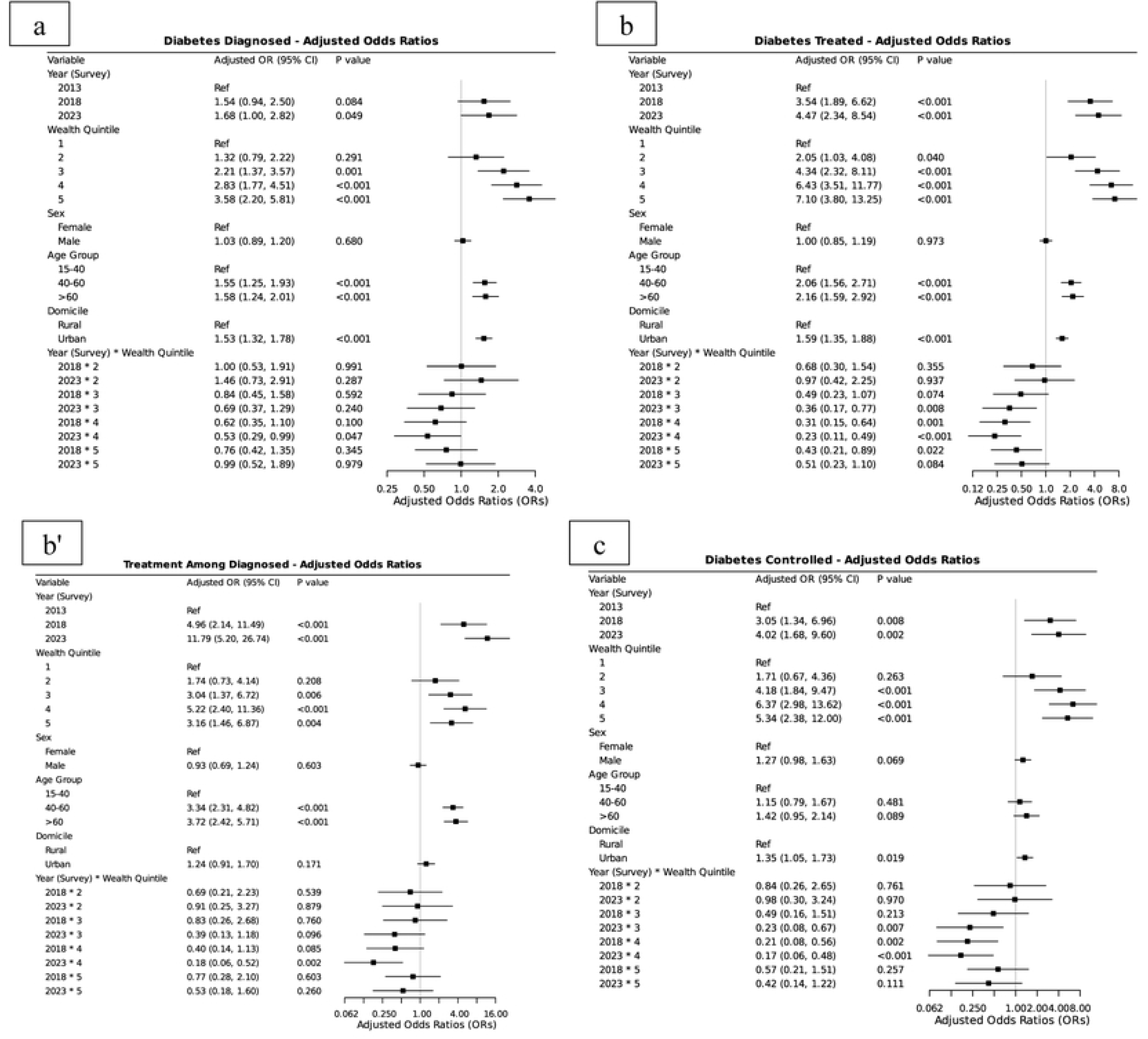
Forest Plot of Adjusted Odds Ratios for Diabetes Care Cascade (a) Diagnosed, (b) Treated, (b’) Treated by Diagnosed, and (c) Controlled. Forest plot of adjusted odds ratios (AORs) with 95% CIs for (a) diagnosis among all with diabetes, (b) treatment among all with diabetes, (b’) treatment among those diagnosed, and (c) glycemic control among all with diabetes Models are survey-weighted logistic regressions adjusting for prespecified covariates (see Methods and Supplementary Table 4 for variable definitions and reference categories).

Among those diagnosed, treatment improves equity. In 2013, only 36.8% (20.3–53.3%) of the poorest (Q1) were treated, compared to 71.2% (62.2–80.1%) in the richest group (Q5). By 2023, treatment coverage had increased markedly and became more equitable across wealth groups, with 89.7% (84.8– 94.6%) of individuals in Q1 and 93.5% (90.4–96.6%) in Q5 receiving treatment (**Figure 3**).

## DISCUSSION

Indonesia has made notable improvements in the proportion of individuals diagnosed and treated for diabetes over the past decade. Still, it is far from meeting the global diabetes targets of 80% of diabetes cases being diagnosed and 80% of diagnosed cases being controlled.^12^ More Indonesians with diabetes are aware of their condition and receiving therapy in 2023 than a decade prior. Yet glycemic control across the diabetes population remained stagnant, and among treated individuals it declined, indicating that gains in coverage have not translated into better outcomes.

### Global Comparison

Compare to regional peers Indonesia’s diagnostic performance lags behind. For example, despite having a higher GDP per capita than India and Bhutan, and higher than Bangladesh’s, Indonesia’s diagnosis rate remains markedly lower, suggesting that economic capacity alone has not translated into diagnostic reach and pointing instead to gaps in case-finding strategy and primary-care implementation. For example, 57.5% of India’s diabetic population is aware of their condition, and 15.2% achieve glycemic control. Bangladesh and Bhutan testing and diagnosis rates are substantially higher (47.2% diagnosed in Bangladesh; 80.5% tested and 62.5% diagnosed in Bhutan).^13–15^ The stagnation in glycemic control we observe mirrors a pattern across many low- and middle-income countries (LMICs), where diabetes awareness and treatment coverage have expanded, yet effective control remains low. ^16^ A cross-country analysis of 28 LMICs found large "leaks" at each cascade step, with fewer than one in four people with diabetes achieving glycemic control and over 75% having unmet care needs. ^16^ Similarly, China saw no significant improvement in control between 2013 and 2018 despite rising diagnosis and treatment, with only about 50% of treated patients adequately controlled in 2018, unchanged from five years earlier. ^17^

### Factors Associated with Care Cascade

Individual-level factors played a significant role across all stages of the diabetes care cascade. Individuals aged 40 and above 60 had higher odds of being diagnosed, treated, and achieving control compared to younger individuals aged 15-40. This trend is consistent with prior studies, including multicountry analyses, which found that older populations are more likely to engage with healthcare systems alongside a greater awareness of diabetes risks.^14,18,19^ The rather poor performance of the diabetes care cascade did not differ by sex. We found that urban-rural disparities exist in diabetes care, with urban residents having higher odds of being diagnosed, treated, and achieving control than rural residents.^20,21^

Urban residents were associated with higher odds of access to diabetes diagnosis and management than rural residents.^14,20–22^ The health workforce and infrastructure are concentrated in cities. For example, Jakarta, as the capital, has over 15 general physicians per 10,000 people, whereas a rural province like West Sulawesi has fewer than 1 per 10,000.^23^ According to the 2019 National Health Facility Research (Rifaskes), while most urban puskesmas (86.7%) can perform blood glucose testing on-site, only 60.9% of remote and very remote puskesmas have this capacity, with 24.5% unable to test at all; HbA1c testing is essentially absent from puskesmas,^24^ insulin cannot be procured or dispensed at the primary level under current regulations,^10^ and primary care provider knowledge of diabetes has been documented to decline over time.^25,26^ Consequently, rural and remote populations are less likely to be screened, diagnosed early, or receive regular monitoring.

The wealth gap was stark: the richest quintile was more likely to be diagnosed, treated, and to have their condition controlled than the poorest. While treatment disparities narrowed, diagnostic gaps persisted, suggesting Indonesia’s primary inequity lies in access to diagnostics, not treatment. This is consistent with JKN: once diagnosed, treatment is broadly and uniformly covered, but case-finding depends on demand-side behavior JKN does not subsidize. Diagnostic inequity likely reflects lower health literacy, reduced screening participation, and limited engagement with health services among lower-income populations. ^27^

### Policy Implication

To expand screening coverage, Indonesia launched a nationwide health screening program in February 2025.^28^ However, the success of this program will depend on addressing several persistent challenges. These include: (1) limited healthcare coverage, (2) inappropriate selection of the population pool, and (3) suboptimal choice of screening tests. In terms of healthcare coverage issues, Indonesia’s screening program has historically been constrained by the service hours of public healthcare facilities, which only accept registrations from 8 am to 1 pm, a schedule that does not match the working population’s needs,^28–29^, and the poor population rarely has access to after-hours private care.^30^

However, there was no mandate requiring working adults to be screened.^31^ The diabetes guideline states that extensive screening is needed for individuals aged 40 or older or those classified as obese (BMI >25). However, those who have a normal BMI (18.5 to 25), despite having lower diabetes prevalence, account for 47% of all undiagnosed diabetes cases (**Supplementary Table 17**). A screening strategy that is too broad may reduce predictive value, but one that is too narrow has risks of missing a substantial portion of at-risk individuals. Indonesia may benefit from adopting a risk-scoring approach, such as those used in Australia, Finland, or Thailand, to identify high-risk individuals more accurately. Test selection is equally consequential.^32–34^ The Ministry of Health’s screening report detected diabetes in only 6% of those screened,^35^ largely reflecting reliance on random blood glucose tests, which are no longer recommended. Future efforts should jointly refine the target population and the diagnostic tests used.

It should be noted that even with expanded screening and increased diagnostic uptake, significant barriers remain in managing patients after diagnosis.**^36,37^** as has been shown in our where glycemic control among treated individuals, from 44.2% (37.2–51.1%) in 2013 to 33.9% (28.2–39.6%) in 2023 (p=0.026), a measurable deterioration in the effectiveness of care among those already reached by the health system, and several mechanisms may explain it. Indonesia’s clinical guidance has been periodically updated by PERKENI — most recently in 2021,**^39^** expanding the role of HbA1c, individualized glycemic targets, and newer agents, and is operationalized at the primary-care level. Prolanis recruitment increased from around 11,000 in 2014 to 250,000 in 2016, and 950,000 members in 2021 (around 300,000 are Diabetes patients), with up to 83.2% active members (revisit after diagnosis).**^40^** Yet this covers only ∼1.5% of the 22 million people with diabetes needing care. Longitudinal Prolanis cohort data and recent provincial evidence show modest, inconsistent HbA1c improvements and declining composite control performance, indicating the program has expanded treatment access without translating into effective control.**^9,41^** Scaling up will require addressing the program’s funding and workforce constraints, including poor access to HbA1c testing for monitoring enrolled patients,**^42^ which can be** addressed through increased financing, improved procurement, and task shifting.**^43^**

## STRENGTHS AND LIMITATIONS

The strength of this study lies in its comprehensive, nationwide analysis of the diabetes care cascade. The larger sample size enables more robust cross-year comparisons than in previous research. It enables in-depth analysis of socioeconomic characteristics and provides detailed trends and factors associated with diabetes care in Indonesia. However, several limitations should be noted. First, the cross-sectional nature of the data limits causal interpretation between individual- and structural-level factors to diabetes care outcomes. Second, the analysis of the control stage is constrained by limited statistical power due to the relatively small number of individuals within this group. Third, variations in data formats across surveys, such as the availability of HbA1c data only in SKI 2023, constrained certain longitudinal analyses.

## CONCLUSIONS

Despite improvements in diagnosis and treatment coverage, the diabetes care cascade in Indonesia remains far from meeting global targets, with glycemic control largely stagnant and socioeconomic disparities persistent across all cascade stages. The 2025 nationwide screening program reflects important political commitment, but its impact will depend on correcting well-documented limitations: inappropriate population targeting, suboptimal test selection, and constrained healthcare coverage. Critically, expanding detection without strengthening downstream chronic disease management risks reproducing the existing pattern of more diagnoses without better outcomes. Addressing both the quality and reach of care simultaneously is essential if Indonesia is to reduce its growing burden of diabetes complications meaningfully.

## Data Availability

The data underlying the findings presented in this study are third-party data owned by the Badan Kebijakan Pembangunan Kesehatan (BKPK), Ministry of Health, Republic of Indonesia. The data were derived from the Riskesdas (Riset Kesehatan Dasar) surveys in 2013, 2018, and 2023. These data are fully available to the public without restriction and can be accessed from the official website of the Pusat Data dan Informasi (PUSDATIN) / Data Repository of the Ministry of Health at the following URL: https://layanandata.kemkes.go.id/katalog-data/riskesdas/ketersediaan-data/riskesdas-2013 https://layanandata.kemkes.go.id/katalog-data/riskesdas/ketersediaan-data/riskesdas-2018 https://layanandata.kemkes.go.id/katalog-data/ski/ketersediaan-data/ski-2023

https://layanandata.kemkes.go.id/katalog-data/riskesdas/ketersediaan-data/riskesdas-2018

https://layanandata.kemkes.go.id/katalog-data/ski/ketersediaan-data/ski-2023

## LIST OF ABBREVIATIONS

BMI: Body Mass Index
CI: Confidence Interval
FPG: Fasting Plasma Glucose
IDF: International Diabetes Federation
NCD: Non-Communicable Diseases
OGTT: Oral Glucose Tolerance Test
AOR: Adjusted Odds Ratio
PSU: Primary Sampling Unit
RISKESDAS: Indonesian Basic Health Research
SES: Socioeconomic Status
SKI: Indonesia Health Survey
SOP: Standard Operating Procedures
WHO: World Health Organization.

## DECLARATIONS

### Consent for publication

Not applicable.

### Availability of data and materials

All data generated or analyzed during this study are included in this published article [and its supplementary information files].

## Funding

This study did not receive any external funding.

## Authors’ contributions

FRM: conceptualization, investigation, data curation, formal analysis, methodology, software, visualization, writing—original draft, and project administration.

RAS: data curation, formal analysis, visualization, and writing—original draft

MQBZ: data curation, formal analysis, visualization, and writing—original draft

AS: conceptualization, investigation, and writing—original draft

GD: Supervisor, data validation, investigation, methodology, validation, and writing—review and editing.

## Acknowledgments

We acknowledge the Health Policy Agency, Ministry of Health, Indonesia, which provided us with the National Health Survey data. We also acknowledge the Indonesia Endowment Fund (LPDP) for supporting the author’s research and study process.

## Declaration of Interest

The authors declare that they have no conflicts of interest.

## STROBE statements

This observational analysis followed STROBE reporting guidelines (checklist in Supplement). Sex-disaggregated analyses were conducted, and interpretation considered sex and gender in line with SAGER guidance.

## SUPPORTING INFORMATION

**Supplementary Text 1. Sampling and Survey Design.** Description of the multistage stratified random sampling methodology used across the 2013 Riskesdas, 2018 Riskesdas, and 2023 SKI national health surveys, including primary sampling unit selection, probability proportional to enrollment size design, and household sampling procedures.

**Supplementary Text 2. Blood Glucose Measurement Protocol.** Description of fasting plasma glucose (FPG), oral glucose tolerance test (OGTT), and HbA1c measurement procedures, including fasting requirements, glucose load administration, and criteria used to classify normal, prediabetes, and diabetes glycaemic status.

**Supplementary Table 1. Response Rate of the Indonesia Health Survey in 2013, 2018, and 2023.** Census block, household, and individual-level response rates for both individual interview and biomedical subsamples across all three survey rounds.

**Supplementary Table 2. Diabetes Category by Lab and History.** Classification matrix of diabetes status combining self-reported diagnosis history with laboratory test results (ADA 2024 criteria), defining undiagnosed, controlled, and uncontrolled diabetes categories.

**Supplementary Table 3. Definition of Diabetes Care Cascade Stages.** Operational definitions for each stage of the diabetes care cascade prevalence, diagnosed, treated, and controlled including denominators, numerators, and reciprocal categories.

**Supplementary Table 4. Operational Definition of Covariates.** Definitions and categorizations of all covariates used in the analysis, including age, sex, domicile, education, occupation, wealth quintile, BMI, tobacco consumption, and insurance status.

**Supplementary Table 5. Characteristics of Interview and Laboratory Samples from Riskesdas 2013, 2018, and SKI 2023.**Unweighted sample sizes and distributions of diabetes status, diagnosis, treatment, age group, sex, and wealth quintile across interview and laboratory subsamples for all three survey years.

**Supplementary Table 6. Comparison of Weighted Prevalence Between Interview, Unadjusted Laboratory, and Adjusted Laboratory Samples.** Post-stratification calibration results comparing weighted prevalence estimates for diabetes status, diagnosis, treatment, age, and sex across interview and laboratory samples before and after raking adjustment.

**Supplementary Table 7. Characteristics of Excluded and Missing Samples from Riskesdas 2013, 2018, and SKI 2023.**Demographic and socioeconomic characteristics of individuals excluded from or missing in the biomedical subsample, including age, sex, domicile, education, occupation, BMI, smoking status, and wealth quintile.

**Supplementary Table 8. Sensitivity Analysis: Glycemic Control Using Alternative Thresholds.** Prevalence of glycemic control estimated using alternative definitions : FPG <140 mg/dL, OGTT <200 mg/dL, combined FPG <126 or OGTT <200, and HbA1c <7% across survey years.

**Supplementary Table 9. Percentage of Diabetes Patients Diagnosed by Subgroup, 2013–2023.** Age-standardized and crude diagnosis rates stratified by sex, age group, BMI, insurance status, wealth quintile, and domicile across all three survey years.

**Supplementary Table 10. Percentage of Diabetes Patients Treated by Subgroup, 2013–2023.** Age-standardized and crude treatment rates stratified by sex, age group, BMI, insurance status, wealth quintile, and domicile across all three survey years.

**Supplementary Table 11. Percentage of Diabetes Patients Controlled by Subgroup, 2013–2023.** Age-standardized and crude glycemic control rates stratified by sex, age group, BMI, insurance status, wealth quintile, and domicile across all three survey years.

**Supplementary Table 12. Percentage of Diagnosed Diabetes Patients Receiving Treatment by Subgroup, 2013–2023.** Age-standardized and crude treatment rates among diagnosed individuals, stratified by sex, age group, BMI, insurance status, wealth quintile, and domicile.

**Supplementary Table 13. Percentage of Treated Diabetes Patients Achieving Glycemic Control by Subgroup, 2013–2023.** Age-standardized and crude glycemic control rates among treated individuals, stratified by sex, age group, BMI, insurance status, wealth quintile, and domicile.

**Supplementary Table 14. Multivariable Logistic Regression: Predictors of Treatment Among Diagnosed Individuals and Glycemic Control Among Treated Individuals.** Adjusted odds ratios and 95% confidence intervals for survey year as a predictor of transition from diagnosis to treatment and from treatment to glycemic control.

**Supplementary Table 15. Wald Test Results for Base Model vs. Interaction Model.** Design-based Wald F-statistics and p-values testing the joint significance of year × wealth quintile interaction terms for diabetes diagnosis, treatment, control, and prevalence outcomes.

**Supplementary Table 16. Adjusted Odds Ratios for Q5 vs. Q1 Wealth Contrast by Survey Year.** Adjusted odds ratios and 95% confidence intervals for the richest (Q5) versus poorest (Q1) wealth quintile contrast across diagnosis, treatment (overall and among diagnosed), and glycemic control outcomes in 2013, 2018, and 2023.

**Supplementary Table 17. BMI Distribution Among Adults With and Without Diabetes Diagnosis.** Proportions of underweight, normal, overweight, and obese BMI categories among diagnosed and undiagnosed adults with diabetes.

**Supplementary Figure 1. Diabetes Care Cascade by Sex and Domicile, Indonesia 2023.** Cascade diagrams showing the proportions of adults with diabetes who are diagnosed, treated, and controlled, stratified by sex (female/male) and domicile (urban/rural) for 2023.

**Supplementary Figure 2. Trends in Diagnosed, Treated, and Controlled Diabetes by Age Group, 2013–2023.** Dot plots showing care cascade coverage rates with 95% confidence intervals for adults aged 15–40, 40–60, and >60 years across the three survey years.

## Notes

### Competing Interest Statement

The authors have declared no competing interest.

### Author Declarations

The data used for this study were obtained from the Riset Kesehatan Dasar (Riskesdas) 2013 and 2018, and the Survei Kesehatan Indonesia (SKI) 2023. These surveys are anonymized, nationally representative datasets collected by the Ministry of Health of the Republic of Indonesia. Ethical clearance for the original surveys was obtained from the National Health Research Ethics Committee of the Ministry of Health. As this study involves secondary analysis of publicly available, non-identifiable data, it was determined to be exempt from the requirement for further Institutional Review Board (IRB) oversight.

